# Prognostic impact of E-wave and A-wave overlap after atrial fibrillation ablation

**DOI:** 10.1101/2024.12.27.24319712

**Authors:** Jumpei Saito, Kato Daiki, Sato Hirotoshi, Toshihiko Matsuda, Yui Koyanagi, Katsuya Yoshihiro, Yuma Gibo, Ishigaki Shigehiro, Soichiro Usumoto, Taro Kimura, Suguru Shimazu, Wataru Igawa, Seitaro Ebara, Toshitaka Okabe, Naoei Isomura, Masahiko Ochiai

## Abstract

**Background:** In adult patients with systolic heart failure, the presence of adjacent, nonoverlapping E and A waves on Doppler echocardiography is associated with optimal cardiac output and favorable clinical outcomes. However, the clinical significance of echocardiographic overlap in patients with atrial fibrillation (AF) remains uncertain. This study aimed to explore the relationship between E-wave and A-wave overlap, assessed the day after catheter ablation, and the recurrence of atrial arrhythmias (AR) following AF ablation.

**Methods:** Patients with AF who underwent first-time arrhythmia ablation were included in this study. Transthoracic echocardiography was performed on the day following catheter ablation to evaluate the presence of E- and A-wave overlap. The relationship between overlap length and recurrence of AR after AF ablation was analyzed.

**Results:** The study included 175 patients (124 males; mean age: 68 years [range 52–79]; mean CHA2DS2-Vasc score: 2 [range 0–4]; and 93 with paroxysmal AF) who underwent AF ablation. There were no significant differences between the two groups in terms of heart failure history or echocardiographic parameters prior to catheter ablation. However, the absolute overlap length was significantly prolonged in the AR group (59 ms [range 9–160] vs. 120 ms [range 28.6–226]; P < .001). Furthermore, the rate of AR was significantly lower in the group without prolonged overlap length (HR, 0.15 [95% CI, 0.07–0.30]; P < .001). These findings were consistent across all AF types.

**Conclusions:** The length of E-wave and A-wave overlap appears to be a significant predictor of AR following AF ablation.

## 1 Introduction

Catheter ablation (CA) remains the cornerstone treatment for atrial fibrillation (AF)^1–5^. However, post-CA recurrence continues to be a significant clinical challenge.^6,7^ The success rate of a single procedure at 12 months varies between 52% and 88%, depending on the type of AF (persistent vs. paroxysmal) and patient characteristics. Most arrhythmic recurrences (AR) occur within the first year following ablation.

Transmitral Doppler echocardiography is typically used to assess cardiac function and identify the E wave, representing the rapid ventricular filling phase, and the A wave, which corresponds to atrial systole. The overlap of these two waves, commonly observed in patients with sinus tachycardia, has been previously reported as an indicator of inadequate diastolic filling time, ultimately leading to reduced cardiac output (Figure 1A)^8^. Conversely, in patients with an excessively low heart rate, the E and A waves remain distinctly separated, resulting in prolonged filling time (Figure 1B)^8^. While this may increase cardiac output during each cycle, the overall cardiac output per minute may be reduced.

**Figure 1.**
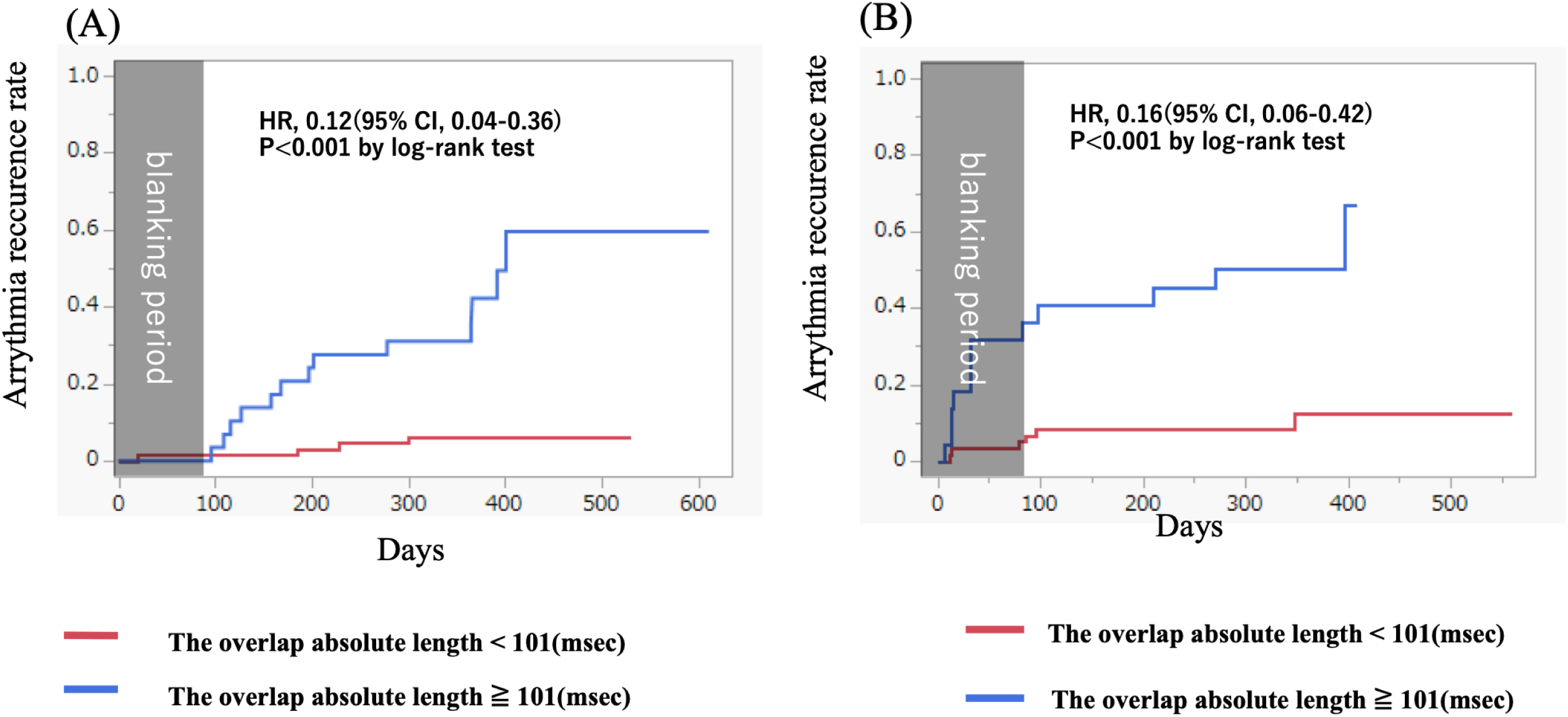
Trans-mitral Doppler echocardiography in patients exhibiting an evident overlap between the E-wave and A-wave, attributable to a relatively elevated heart rate (**A**). Conversely, in patients displaying no such overlap, the heart rate is relatively lower (**B**). An evident wave overlap indicates inadequate left ventricular filling time, which results in reduced cardiac output. Conversely, an evident wave separation indicates sufficient left ventricular filling time but also results in reduced cardiac output due to a lower stroke volume.

In adult patients with systolic heart failure, the presence of adjacent, nonoverlapping E and A waves on Doppler echocardiography during sinus rhythm has been associated with maximal cardiac output and favorable clinical outcomes^9,10^. However, the clinical significance of echocardiographic overlap in patients undergoing ablation for atrial fibrillation remains unclear. This study aimed to investigate the relationship between E-wave and A-wave overlap immediately following catheter ablation and the recurrence of atrial arrhythmias after AF ablation.

## 2 Methods

## 3 Patients

We included 175 patients who underwent their first AF ablation procedure at our hospital between January 2022 and June 2023. The exclusion criteria were as follows: age < 20 years, implanted pacemaker, and recurrent AF on the day after catheter ablation. The study was conducted in accordance with the principles of the Declaration of Helsinki. Written informed consent for the ablation procedure and participation in the study was obtained from all patients, and the study protocol was approved by our Institutional Review Board (ID: 2024-184-A).

## 4 Catheter Settings and Ablation Protocol

AF ablation was performed under deep sedation using propofol and fentanyl. A 20-polar electrode catheter (BeeAT; Japan Lifeline, Tokyo, Japan) was introduced into the coronary sinus via the internal jugular vein. Two or three long sheaths were inserted into the left atrium through the right femoral vein. Pulmonary vein isolation was achieved using either a radiofrequency catheter or a cryoballoon (Arctic Front; Medtronic, Minneapolis, MN, USA) with the guidance of a three-dimensional mapping system (CARTO [Biosense Webster, Diamond Bar, CA, USA] or Ensite NavX [Abbott, Chicago, IL, USA]).

Additional ablation strategies, such as cavotricuspid isthmus ablation, superior vena cava isolation, left atrial linear ablations, and other substrate ablations, were performed at the discretion of the attending physician or operator.

## 5 Echocardiographic Assessment

The patient underwent transthoracic echocardiography on the day following catheter ablation for AF. Transthoracic echocardiography was performed according to the current American Society of Echocardiography guidelines by experienced sonographers who were blinded to the study protocol. Heart rate was measured simultaneously using a three-lead electrocardiogram. Left ventricular ejection fraction (LVEF) was calculated using the modified Simpson method. E-wave deceleration time was measured using pulse Doppler echocardiography at the transmitral flow in the apical four-chamber view. In transmitral flow, the overlap between the E- and A-waves was measured. If there was no overlap, the distance between the two waves was expressed as negative (Figure 1).

## 6 Follow Up

The patient underwent transthoracic echocardiography on the day following catheter ablation for AF. Patients were scheduled to visit the outpatient clinic at 1, 3, 6, and 12 months after ablation, and annually thereafter. A 24-hour Holter ECG was performed at 3, 6, and 12 months. BNP levels were measured one month after ablation, and sinus rhythm was confirmed using a 12-lead ECG at the same time. Three months after AF ablation, all patients wore a 24-hour Holter ECG to confirm sinus rhythm, and BNP levels were measured. The occurrence of atrial fibrillation, atrial flutter, and atrial tachycardia lasting > 30 s after a 3-month blanking period was defined as AR. Early recurrence was defined as recurrence within the first three months.

## 7 Statistical Analysis

Continuous data are expressed as medians with interquartile ranges, while categorical data are presented as absolute values and percentages. Differences in variables between the blocked and reconnected segments were analyzed using the Wilcoxon signed-rank test for continuous variables and the χ² test or Fisher’s exact test for categorical variables. All statistical analyses were performed using commercial software (JMP).

The discriminatory accuracy of the absolute overlap length for predicting AR was evaluated using the area under the curve (AUC) of the receiver operating characteristic (ROC) curve, derived from logistic regression analysis. The cut-off value for the absolute overlap length that optimized the balance between sensitivity and specificity for AR was determined as the point on the ROC curve closest to the upper left-hand corner of the AUC.

Patients were then stratified into two groups based on the previously identified cut-off values for absolute overlap length. AR rates were assessed using the Kaplan-Meier method and compared between groups using the log-rank test. Statistical significance was defined as P < .05.

## 8 Results

## 9 Patient Characteristics

The baseline characteristics of the patients are summarized in Table 1. The mean age was 68 years; 124 patients (70.9%) were male and 93 patients (53.1%) had paroxysmal AF. No significant differences in the history of heart failure or echocardiographic parameters before CA were observed between the two groups.

**Table 1.** Baseline patient characteristics and echocardiogram parameter.

| Characteristics | Non recurrence (N=139) | Recurrence (N=36) | P value |
| --- | --- | --- | --- |
| age, y | 68 (52,79) | 68 (51,80) | 0.67 |
| male sex, n (%) | 97 (69.8) | 27 (75) | 0.53 |
| body mass index, kg/m<sup>2</sup> | 23.9 (19.4, 29.0) | 24.4 (20.1, 32.2) | 0.21 |
| CHAD<sup>2</sup>DS<sup>2</sup>-VASc score | 2 (0, 4) | 2 (0, 4.3) | 0.72 |
| paroxysmal AF, n (%) | 76 (54.7) | 17 (47.2) | 0.42 |
| heart failure, n (%) | 24 (17.3) | 8(23.5) | 0.41 |
| **echocardiogram before ablation** |  |  |  |
| LVEF, % | 59 (43.4, 67) | 58 (41.2, 68.4) | 0.53 |
| LAVI, ml/m<sup>3</sup> | 37 (23, 53.7) | 38.5 (20.5, 64) | 0.10 |

Data are presented as n (%) or median (interquartile range).
Abbreviations: CHA<sup>2</sup>DS<sup>2</sup>-VASc, congestive heart failure, hypertension, age $\geq 75$ years (doubled), diabetes mellitus, prior stroke or transient ischemic attack or thromboembolism (doubled), vascular disease, age 65–74 years, AF, atrial fibrillation, LVEF, left ventricular ejection fraction, LAVI, left atrial volume index

## 10 E-Wave and A-Wave Overlap Length

When the E and A-waves overlapped, their lengths were analyzed as they were; when the E and A-waves were far apart, the length of the distance between them was expressed as a negative sign, but this negative sign was omitted, and the analysis was carried out using absolute values. The absolute overlap length was significantly prolonged in the AR group (Table 2). When the two groups were further divided into two groups, with and without overlap, the overlap length was significantly prolonged in the AR group.

**Table 2.** Echocardiographic Parameters after AF ablation.

|  | Non recurrence (N=139) | Recurrence (N=36) | P value |
| --- | --- | --- | --- |
| LVEF, % | 60 (46, 66) | 61 (47.7, 66.3) | 0.54 |
| LAVI, ml/m<sup>3</sup> | 38 (23, 57) | 37.5 (26.7, 70.8) | 0.22 |
| E-wave velocity, cm/sec | 77.9 (57.4, 110.1) | 82.4 (55.5, 118.8) | 0.72 |
| A-wave velocity, cm/sec | 54.4 (31.5, 83.3) | 50.4 (22.8, 90.7) | 0.30 |
| E/A ratio | 1.48 (0.89, 2.71) | 1.60 (0.85, 3.82) | 0.02 |
| E/A overlap absolute length, msec | 59 (9, 160) | 120 (28.6, 226) | < 0.001 |
| heart rate during echocardiography, bpm | 74 (62, 89) | 73 (60, 89) | 0.32 |

|  | non recurrence (N=49) | recurrence (N=21) |  |
| --- | --- | --- | --- |
| E/A overlap length, msec | 77 (22, 160) | 121 (23.8, 170.8) | 0.03 |
|  | non recurrence (N=90) | recurrence (N=15) |  |
| E/A non-overlap length, msec | -46.5 (-120, -5.3) | -108 (-264.4, -24.4) | < 0.001 |

Data are presented as the median (interquartile range).
Abbreviations: AF, atrial fibrillation; LVEF, left ventricular ejection fraction; LAVI, left atrial volume index; E/A, E-wave and A-wave

ROC curve analysis showed that the area under the curve (AUC) for the absolute overlap length was 0.760 (Figure 2A). The cutoff value was 101 ms (sensitivity, 0.722; specificity, 0.813). When the two groups were further divided into two groups, with and without overlap, the AUC for the overlap length was also high (Figure 2B and 2C). The overlap group and non-overlap group, the optimal cut-off value of the overlap length was approximately 101 (sensitivity, 0.801; specificity, 0.710) and -104 msec (sensitivity, 0.600; specificity, 0.878).

**Figure 2.**
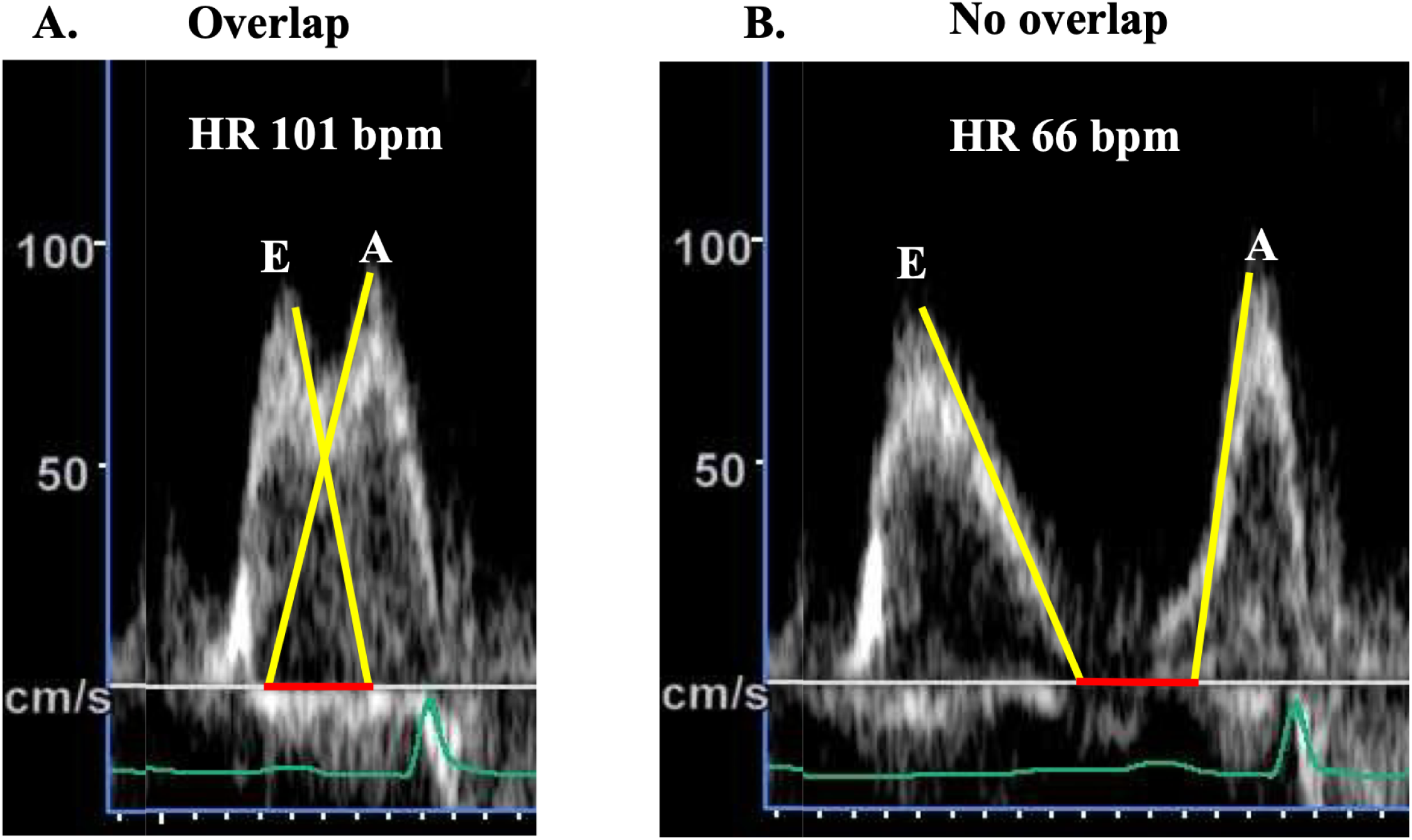
(**A**) AUC for the overlap absolute length in all patients was 0.760. (**B**) AUC for the overlap length in E and A wave overlap group was 0.700. (**C**) AUC for the overlap length in E and A wave non-overlap group was 0.775. AUC, area under the curve

Kaplan-Meier curves showed that patients with an overlap absolute length of > 101 ms had a significantly higher incidence of AR than those without (Figure 3A). In the overlapping and non-overlapping groups, patients with a prolonged overlap also had a significantly higher incidence of AR than those without a prolonged overlap (Figure 3B and 3C). The results were consistent regardless of AF type (Figure 4).

**Figure 3.**
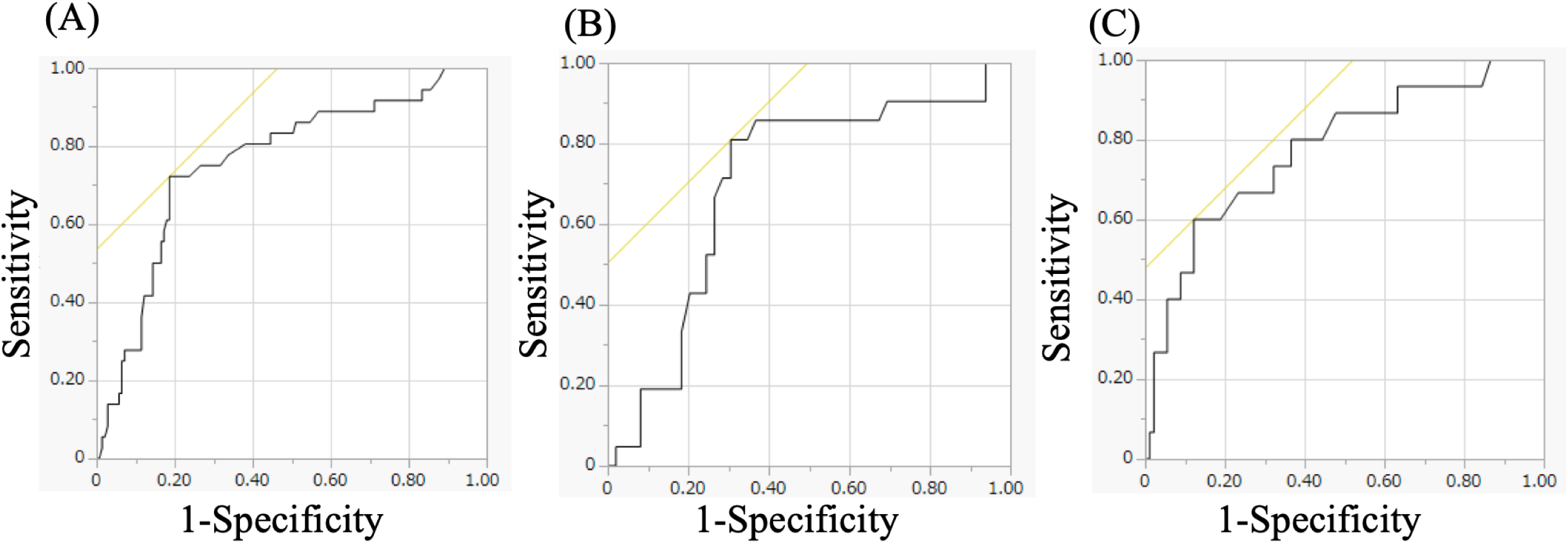
(**A**) Kaplan–Meier curve showing the arrhythmia recurrence rate for patients with (blue) and without (red) prolonged the overlap absolute length after AF ablation. (**B**) Kaplan–Meier curve showing the arrhythmia recurrence rate for patients with (blue) and without (red) prolonged the overlap length in E and A wave overlap group after AF ablation. (**C**) Kaplan–Meier curve showing the arrhythmia recurrence rate for patients with (blue) and without (red) prolonged the overlap length in E and A wave non-overlap group after AF ablation. AF; atrial fibrillation

**Figure 4.**
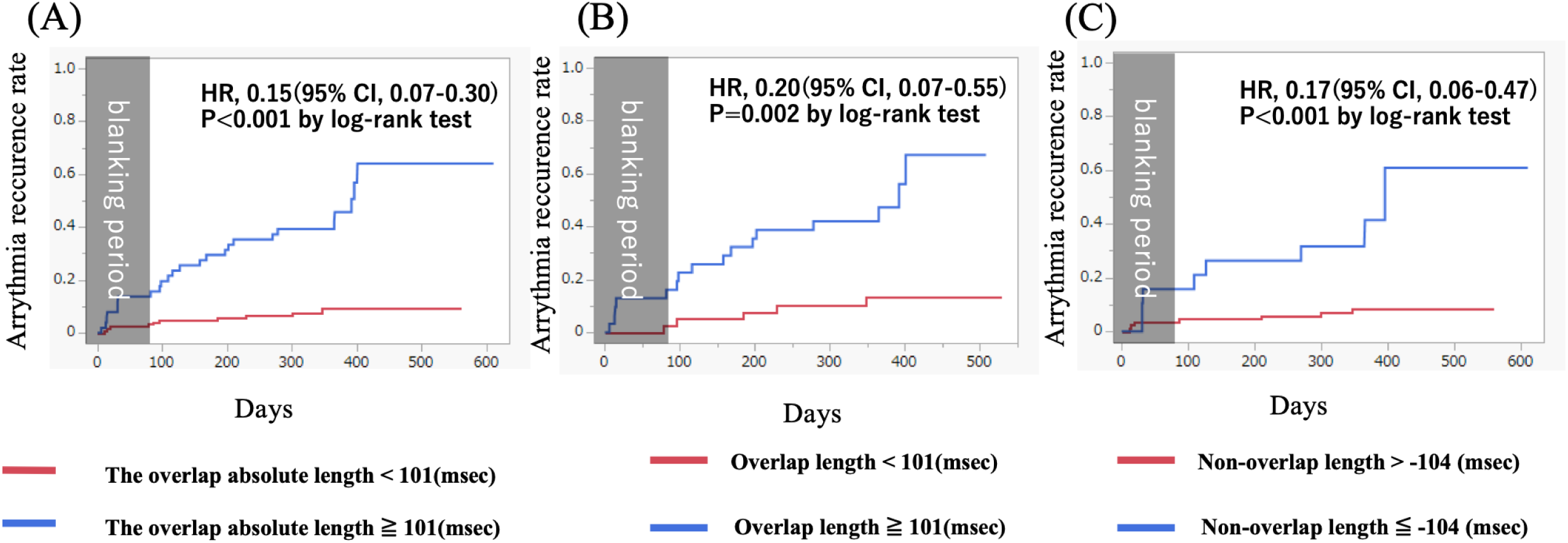
(**A**) Kaplan–Meier curve showing the arrhythmia recurrence rate for patients with (blue) and without (red) prolonged the overlap absolute length in paroxysmal AF patients. (**B**) Kaplan–Meier curve showing the arrhythmia recurrence rate for patients with (blue) and without (red) prolonged the overlap absolute length in non-paroxysmal AF patients. AF; atrial fibrillation

## 11 Discussion

### Main Finding

The present study is an inaugural investigation to evaluate the association of E-wave and A-wave overlap lengths and AR after AF ablation. The main findings are summarized as follows:

1. The E-wave and A-wave overlap lengths are associated with an increased risk of AR.
2. When the data were divided into two groups–one comprising cases with an overlap and the other comprising cases without an overlap–the overlap length was similarly associated with an increased risk of AR.
3. When the data were divided into two groups, one comprising patients with paroxysmal AF and the other comprising patients with non-paroxysmal AF, the overlap length was similarly associated with an increased risk of AR.

### Importance of Left Atrial Function

LA remodeling is important for the development and persistence of AF. This remodeling process encompasses three principal domains: electrical, contractile, and structural remodeling^11^, and there are various evaluation methods for assessing left atrial function, including echocardiography, computed tomography (CT), and magnetic resonance imaging (MRI). Previous studies have shown that impaired left atrial function on echocardiography may be a factor in the development of atrial fibrillation in the general population^12–14^. It has been also reported that patients with AF were impaired left atrial function, and impaired atrial function was the risk of recurrence after AF ablation and clinical outcome^15–21^. Therefore, to enhance patient selection and increase the success rate of AF ablation, a more precise understanding of LA remodeling in AF is crucial. This facilitated optimal treatment planning and risk stratification. However, our study did not assess left atrial strain or evaluate left atrial function using CT or MRI.

### E-Wave and A-Wave Overlap Length

Pulsed-wave transmitral flow Doppler echocardiography may prove to be an effective means of evaluating the relationship between the heart rate and left ventricular filling. During sinus tachycardia, E-waves and A-wave merge^22^. In a cohort of healthy individuals, a merged A-wave exhibited an increase in height as the heart rate increased, which might be a compensatory mechanism for reduced left ventricular filling^23^. However, such compensation may not be effective in patients with AF because of impaired left atrial function. There are numerous indices of left atrial function, and echocardiographic parameters alone include the left atrial volumetric index, septal and lateral late diastolic peak tissue Doppler velocity, and left atrial strain parameters. Angelini et al. reported that echocardiographic parameters on the day after AF ablation were predictive of AF recurrence, a finding analogous to our study^24^. However, since E-wave and A-wave overlap lengths were not evaluated, we reported that the overlap length is a predictor of AF recurrence. Heart rates in the recurrent and non-recurrent groups did not differ significantly. Our study indicated that the overlap length was more likely to be prolonged in the recurrence group at similar heart rates. Therefore, a longer overlap length may indicate impaired left atrial function. We believe that this is a useful approach because it provides one of the most straightforward echocardiographic parameters for measurement.

## 12 Limitation

This study had some limitations. First, the single-center, relatively small study population may have affected the reliability of the statistical findings. Thus, large-scale multicenter studies are required to validate this hypothesis. Second, it was not possible to accurately identify asymptomatic atrial arrhythmias outside of the time when the patients wore a 24-hour Holter ECG. Third, this study lacked longitudinal observations, and the long-term outcomes are unknown. In addition, it is unclear whether echocardiographic parameters in the chronic phase affect outcomes.

## 13 Conclusions

E and A-wave overlap length might be a significant predictor for AR.

## Data availability statement

The original contributions presented in the study are included in the article, and further inquiries can be directed to the corresponding author.

## Ethics approval statement

This study, which involved human participants, was approved by the Showa University Ethics Committee (ID: 2024-184-A).

## Patient consent statement

The patients provided written informed consent to participate in this study.

